# Epidemiology of myocarditis and pericarditis following mRNA vaccines in Ontario, Canada: by vaccine product, schedule and interval

**DOI:** 10.1101/2021.12.02.21267156

**Authors:** Sarah A. Buchan, Chi Yon Seo, Caitlin Johnson, Sarah Alley, Jeffrey C. Kwong, Sharifa Nasreen, Andrew Calzavara, Diane Lu, Tara M. Harris, Kelly Yu, Sarah E. Wilson

**Affiliations:** Public Health Ontario, ON, Canada; Dalla Lana School of Public Health, University of Toronto, Toronto, ON, Canada; ICES, Toronto, ON, Canada; Department of Family and Community Medicine, University of Toronto, Toronto, ON, Canada; University Health Network, Toronto, ON, Canada

## Abstract

**Importance:** Increased rates of myocarditis/pericarditis following COVID-19 mRNA vaccines have been observed. However, little data are available related to product-specific differences, which have important programmatic impacts.

**Objective:** The objective of this study was to estimate reporting rates of myocarditis/pericarditis following COVID-19 mRNA vaccine by product, age, sex, and dose number, as well inter-dose interval.

**Design:** We conducted a population-based cohort study using passive vaccine safety surveillance data. All individuals in Ontario, Canada who received at least one dose of COVID-19 mRNA vaccine between December 14, 2020 and September 4, 2021 were included.

**Setting:** This study was conducted in Ontario, Canada (population: 14.7 million) using the provincial COVID-19 vaccine registry and provincial adverse events following immunization database.

**Participants:** We included all individuals with a reported episode of myocarditis/pericarditis following COVID-19 vaccine in the study period. We obtained information on all doses administered in the province to calculate reporting rates.

**Exposure:** Receipt of COVID-19 mRNA vaccine (mRNA-1273 [Moderna Spikevax] or BNT162b2 [Pfizer-BioNTech Comirnaty]).

**Main Outcome(s) and Measure(s):** Reported rate of myocarditis/pericarditis meeting level 1-3 of the Brighton Collaboration case definitions.

**Results:** There were 19,740,741 doses of mRNA vaccines administered and 297 reports of myocarditis/pericarditis meeting our inclusion criteria. Among these, 69.7% occurred following the second dose of COVID-19 mRNA vaccine and 76.8% occurred in males. The median age of individuals with a reported event was 24 years. The highest reporting rate of myocarditis/pericarditis was observed in males aged 18-24 years following mRNA-1273 as the second dose; the rate in this age group was 5.1 (95% CI 1.9-15.5) times higher than the rate following BNT162b2 as the second dose. Overall reporting rates were higher when the inter-dose interval was shorter (i.e., ≤30 days) for both vaccine products. Among individuals who received mRNA-1273 for the second dose, rates were higher for those who had a heterologous as opposed to homologous vaccine schedule.

**Conclusions and Relevance:** Our results suggest that vaccine product, inter-dose interval and vaccine schedule combinations may play a role in the risk of myocarditis/pericarditis, in addition to age and sex. Certain programmatic strategies could reduce the risk of myocarditis/pericarditis following mRNA vaccines.

## Introduction

Post-marketing vaccine safety surveillance systems in multiple countries have identified a likely association between myocarditis and pericarditis following BNT162b2 (Pfizer-BioNTech Comirnaty) and mRNA-1273 (Moderna Spikevax) COVID-19 mRNA vaccines.^1-5^ In Ontario, Canada, (population approximately 14.7 million), enhanced surveillance for myocarditis/pericarditis following mRNA vaccines began in early June 2021. This consisted of healthcare provider communication from the Ontario Ministry of Health and Public Health Ontario, and hospital-led algorithms for clinical investigations and management that also included instructions on reporting events to Ontario’s existing passive vaccine safety surveillance system. This enhanced surveillance directive (ESD) coincided with a number of changes to Ontario’s COVID-19 vaccine program including: expanded vaccine eligibility to young adults and adolescents (Health Canada authorization of BNT162b2 for individuals aged 12-15 years occurred on May 5, 2021), a large acceleration in vaccine supply and administration of second doses to the population, permissive language from Canada’s National Advisory Committee on Immunization on the use of heterologous mRNA vaccine schedules,^6^ and over the course of the summer of 2021, a gradual return to the scheduling of second doses in accordance with the product monograph (PM) interval following a period of extended intervals between dose 1 and 2 (hereafter referred to as inter-dose interval) to maximize the number of individuals protected with a first dose of vaccine.^6^ These programmatic changes provided an opportunity to examine the risk of myocarditis/pericarditis in relation to a number of factors.

Our objectives were to examine reporting rates of myocarditis/pericarditis following mRNA vaccines by age, sex, vaccine product, dose number, inter-dose interval and homologous/heterologous vaccine schedule, using passive vaccine safety surveillance data.

## Methods

We used the Public Health Case and Contact Management Solution (CCM), Ontario’s electronic reporting system for COVID-19 adverse events following immunization (AEFI), to identify reports of myocarditis and pericarditis following a COVID-19 vaccine reported between December 14, 2020 (the start of Ontario’s immunization program) and September 4, 2021. In Ontario, reporting of AEFI by healthcare providers is mandated by provincial public health legislation; voluntary reporting by vaccine recipients or their caregivers also occurs.^7^ Reports are submitted to local Public Health Units (PHUs) where additional investigation of the event occurs to obtain supporting information (e.g., laboratory findings, diagnostic imaging).

Events were identified through both a keyword search (i.e., ‘myocarditis’ or ‘pericarditis’) and where cardiovascular injury or myocarditis/pericarditis was selected from a list of pre-defined adverse events. Case level review of all reports was completed by a group of specialized nurses and physicians on the PHO vaccine safety team to assign a level of diagnostic certainty using Brighton Collaboration (BC) case definitions for myocarditis or pericarditis, as appropriate.^8^ We restricted our analyses to events meeting BC levels 1-3 of diagnostic certainty. In sensitivity analyses that examined only myocarditis, AEFI reports with physician diagnoses of myopericarditis and perimyocarditis were included only if the BC case definition (levels 1-2) for myocarditis was met. We included all reports following vaccination, regardless of time since vaccination, in crude reporting rates. We used a 7-day risk interval in analyses of observed versus expected number of events.

To calculate reporting rates, we extracted information from the Ontario Ministry of Health’s COVaxON database, the provincial COVID-19 vaccine registry. We calculated reporting rates of myocarditis/ pericarditis combined per million doses of COVID-19 mRNA vaccine by age, sex, dose number and vaccine product. Confidence intervals were calculated using the Poisson exact method. Our primary analysis was restricted to individuals who initiated their vaccine series on or after June 1, 2021, in order to account for any increase in AEFI reporting following the increased awareness resulting from media reports and the provincial ESD for myocarditis/pericarditis that began in early June 2021. This timing also coincided with other changes to the vaccine program, including implementation of heterologous mRNA schedules (Supplementary Figure 1). We also examined reporting rates of heterologous or homologous vaccine schedules, and by inter-dose interval, for the total population and in males aged 18-24 years, restricted to individuals who received dose 2 (regardless of dose 1 date) on or after June 1, 2021 in order to maximize our sample of dose 2 recipients during the period of enhanced surveillance. We selected the interval groupings by examining the distribution of intervals among individuals receiving a second dose and to align with the product monograph(s) and programmatic decisions (i.e., extended inter-dose intervals). In order to estimate an overall rate following dose 2 by product, we used Poisson regression and adjusted for dose 1 product and interval.

We also performed the analysis for myocarditis/pericarditis outcomes for the entire reporting period (i.e., any dose received December 14, 2020 – September 4, 2021).

Finally, we compared our observed events of myocarditis/pericarditis to those expected based on historical data, following the methodology outlined by Mahaux et al.^9^ Historical background rates from the Ontario population were obtained from linked health administrative databases. These rates reflected episodes of myocarditis/pericarditis (see Supplementary Table 1 for ICD-10-CA definitions) obtained from the Canadian Institute for Health Information’s Discharge Abstract Database and National Ambulatory Care Reporting System, reflecting hospitalizations and emergency department visits, respectively. These databases were linked using unique encoded identifiers and analyzed at ICES (formerly the Institute for Clinical Evaluative Sciences). We used the mean rate from 2015-2019 to calculate expected events by age and sex based on the number of vaccines that were administered in each group, using a 7-day risk interval, chosen because the majority of events occurred within this time frame. We estimated a range in expected events by using the confidence limits of the background rates. We focused this part of the analysis on events occurring in individuals who received their second dose on or after June 1, 2021.

The Public Health Ontario Ethics Review Board determined that this project did not require research ethics committee approval as the activities described in this manuscript were conducted in fulfillment of Public Health Ontario’s legislated mandate “to provide scientific and technical advice and support to the health care system and the Government of Ontario in order to protect and promote the health of Ontarians” (Ontario Agency for Health Protection and Promotion Act, SO 2007, c 10) and are therefore considered public health practice, not research.

## Results

Between December 14, 2020 and September 4, 2021, there were 19,740,741 doses of mRNA vaccines administered in Ontario and 417 reports of myocarditis or pericarditis reported to the provincial AEFI system. Of these, 297 met the inclusion criteria based on the BC case definitions (level 1-3); among these, 69.7% occurred following the second dose of COVID-19 mRNA vaccine, 76.8% occurred in males, and the median age of individuals with a reported event was 24 years (Table 1). Events were classified as myopericarditis (36.0%), followed by myocarditis (35.4%), and pericariditis (28.6%). Nearly all (97.6%) events involved an emergency department visit, with 70.7% of events also leading to a hospital admission. The proportion of individuals hospitalized was 82.9%, 38.8%, and 84.1% for myocarditis, pericarditis, and myopericarditis, respectively (Supplementary Table 2). The median time to onset was three days after vaccine administration (interquartile range: 2-8; range: 0-73). Most events (73.9%) with a known onset date occurred within 7 days of vaccine administration. For events following dose 2, 86.9% occurred within 7 days of vaccine with 97.1% occurring within 30 days (Supplementary Figure 2).

**Table 1.** Characteristics of myocarditis/pericarditis reports following COVID-19 mRNA vaccines

|  | After dose 1 (N=90) |  | After dose 2 (N=207) |  | Total (N=297) |
| --- | --- | --- | --- | --- | --- |
|  | Dose administered before June 1 | Dose administered on or After June 1 | Dose administered before June 1 | Dose administered on or After June 1 |  |
| **Total number of reports** | 50 | 40 | 5 | 202 | 297 |
| **Median age, years (range)** | 32 (12 – 81) | 23 (13 – 76) | 50 (34 – 61) | 23 (12 – 81) | 24 (12 – 81) |
| **Age group (years)** |  |  |  |  |  |
| 12-17 | 5 (10.0%) | 14 (35.0%) | 0 (0.0%) | 36 (17.8%) | 55 (18.5%) |
| 18-24 | 12 (24.0%) | 7 (17.5%) | 0 (0.0%) | 77 (38.1%) | 96 (32.3%) |
| 25-39 | 11 (22.0%) | 10 (25.0%) | 2 (40.0%) | 49 (24.3%) | 72 (24.2%) |
| ≥40 | 22 (44.0%) | 9 (22.5%) | 3 (60.0%) | 40 (19.8%) | 74 (24.9%) |
| **Sex** |  |  |  |  |  |
| Male | 32 (64.0%) | 30 (75.0%) | 2 (40.0%) | 164 (81.2%) | 228 (76.8%) |
| Female | 18 (36.0%) | 10 (25.0%) | 3 (60.0%) | 38 (18.8%) | 69 (23.2%) |
| **Median time to onset, days (interquartile range)\*** | 14.5 (7-29) | 4 (2-14) | 2 (2 – 73) | 2 (1-3) | 3 (2-8) |
| **Vaccine product** |  |  |  |  |  |
| BNT162b2 | 39 (78.0%) | 29 (72.5%) | 4 (80.0%) | 87 (43.1%) | 159 (53.5%) |
| mRNA-1273 | 11 (22.0%) | 11 (27.5%) | 1 (20.0%) | 115 (56.9%) | 138 (46.5%) |
| **Clinical diagnosis** |  |  |  |  |  |
| Myocarditis | 18 (36.0%) | 13 (32.5%) | 2 (40.0%) | 72 (35.6%) | 105 (35.4%) |
| Pericarditis | 23 (46.0%) | 15 (37.5%) | 2 (40.0%) | 45 (22.3%) | 85 (28.6%) |
| Myopericarditis** | 9 (18.0%) | 12 (30.0%) | 1 (20.0%) | 85 (42.1%) | 107 (36.0%) |
| **Healthcare utilization/outcome** |  |  |  |  |  |
| Emergency department visit | 49 (98.0%) | 37 (92.5%) | 5 (100.0%) | 199 (98.5%) | 290 (97.6%) |
| In-patient hospitalization | 32 (64.0%) | 24 (60.0%) | 4 (80.0%) | 150 (74.3%) | 210 (70.7%) |
| Intensive care unit admission | 1 (2.0%) | 3 (7.5%) | 0 (0.0%) | 10 (5.0%) | 14 (4.7%) |
| Death | 0 | 0 | 0 | 0 | 0 |

\*2 reports with unknown time to onset were excluded from this calculation.
\*\*Includes “myocarditis/pericarditis” (n=2), myopericarditis (n=81), and peri myocarditis (n=24).

In our primary analysis focusing on those who initiated their vaccination series on or after June 1, 2021, the reporting rate of myocarditis or pericarditis was higher following the second dose of mRNA vaccine than after the first dose, particularly for those individuals receiving mRNA-1273 as the second dose of the series (Table 2). The highest reporting rate of myocarditis or pericarditis was observed in males aged 18-24 years following mRNA-1273 as the second dose, which (in our primary analysis) was 5.1 (95% CI 1.9-15.5) times higher than the rate following BNT162b2 as the second dose (299.5 vs. 59.2 per million doses, respectively). The second highest reporting rate was observed among males 12-17 following their second dose of BNT162b2 (97.3 per million [95% CI 60.3-148.8]). However, confidence intervals were wide. In a sensitivity analysis restricting the analysis to those meeting level 1 or 2 of the BC case definition for myocarditis only, our observed patterns remained unchanged (Supplementary Table 3). When we performed the analysis for myocarditis/pericarditis outcomes for the entire reporting period, the results were similar (Supplementary Table 4; rates following dose 2 by age in years and product in Supplementary Figure 3).

**Table 2.** Crude reporting rate of myocarditis/pericarditis per million doses administered by vaccine product, dose number, age, and sex: series initiation on or after June 1, 2021

| **BNT162b2** |  |  |  |  |  |  |
| --- | --- | --- | --- | --- | --- | --- |
|  | All |  | Female |  | Male |  |
| **Age group (years)** | Dose 1 | Dose 2 | Dose 1 | Dose 2 | Dose 1 | Dose 2 |
| **12-17\*** | 27.3 (14.9 - 45.8) | 54.4 (34.5 - 81.7) | 20.1 (6.5 - 47.0) | 9.7 (1.2 - 35.1) | 34.2 (15.6 - 64.9) | 97.3 (60.3 - 148.8) |
| **18-24** | 17.9 (5.8 - 41.7) | 44.3 (17.8 - 91.3) | 7.9 (0.2 - 44.1) | 27.4 (3.3 - 99.0) | 26.2 (7.1 - 67.0) | 59.2 (19.2 - 138.1) |
| **25-39** | 13.0 (5.2 - 26.8) | 16.0 (5.2 - 37.4) | 3.9 (0.1 - 21.6) | 19.7 (4.1 - 57.6) | 21.5 (7.9 - 46.7) | 12.6 (1.5 - 45.4) |
| **≥40** | 5.9 (1.2 - 17.3) | 0.0 (0.0 - 11.7) | 4.0 (0.1 - 22.3) | 0.0 (0.0 - 23.5) | 7.8 (0.9 - 28.3) | 0.0 (0.0 - 23.3) |
| **Total** | 15.6 (10.4 - 22.4) | 29.0 (20.2 - 40.3) | 8.9 (3.9 - 17.6) | 11.9 (4.8 - 24.5) | 21.8 (13.5 - 33.3) | 45.3 (30.1 - 65.5) |
| **mRNA-1273** |  |  |  |  |  |  |
|  | All |  | Female |  | Male |  |
|  | Dose 1 | Dose 2 | Dose 1 | Dose 2 | Dose 1 | Dose 2 |
| **12-17\*** | - | - | - | - | - | - |
| **18-24** | 21.6 (2.6 - 77.9) | 195.5 (117.7 - 305.3) | 0.0 (0.0 - 95.1) | 69.1 (14.2 - 201.9) | 37.2 (4.5 - 134.6) | 299.5 (171.2 - 486.4) |
| **25-39** | 16.2 (3.3 - 47.3) | 58.7 (30.3 - 102.6) | 0.0 (0.0 - 45.4) | 21.5 (2.6 - 77.7) | 28.8 (5.9 - 84.3) | 90.1 (43.2 - 165.7) |
| **≥40** | 30.0 (11.0 - 65.2) | 0.0 (0.0 - 19.0) | 22.0 (2.7 - 79.4) | 0.0 (0.0 - 40.9) | 36.7 (10.0 - 93.9) | 0.0 (0.0 - 35.6) |
| **Total** | 23.0 (11.5 - 41.1) | 62.5 (42.4 - 88.6) | 9.5 (1.1 - 34.2) | 22.0 (7.1 - 51.4) | 33.7 (15.4 - 64.0) | 96.8 (63.2 - 141.9) |

\*Estimates were not provided for individuals aged 12-17 for mRNA-1273 because this product was not used for this age group in Ontario.

To explore differences in the rate of myocarditis/pericarditis following the second dose of mRNA-1273 vs. BNT162b2, we also examined rates by mixed schedule and inter-dose interval (Figure 1a; further data in Supplementary Table 5). Among all ages and sexes combined, rates of myocarditis/pericarditis were higher for individuals with shorter inter-dose intervals (≤30 days vs. ≥56 days) and the unadjusted rate ratios comparing these intervals were similar for mRNA-1273 (RR= 5.2, 95% CI 2.6-10.0) and BNT162b2 (RR=5.5, 95% CI 3.1-9.6). We also examined overall rates by inter-dose interval within homologous or heterologous schedules (Figure 1b); the highest rate reported was in those who received BNT162b2 followed by mRNA-1273 with an inter-dose interval of ≤30 days. Among males aged 18-24, rates in those who received a second dose of mRNA-1273 (regardless of first dose product) were significantly higher than in those who received two doses of BNT162b2 (Table 3). The rate among males aged 18-24 receiving two doses of mRNA-1273 were lower than those who received BNT162b2 followed by mRNA-1273 (288.4 per million [95% CI 190.1-419.6] vs. 337.6 per million [95% CI 226.1-484.9], respectively). There were no reported events in males aged 18-24 years who received a first dose of mRNA-1273 followed by a second dose of BNT162b2; however fewer than 9,000 males in this age group received this schedule. Within each of these schedules (i.e., BNT162b2-BNT162b2, mRNA-1273-mRNA-1273, BNT162b2-mRNA-1273) rates were lower with a longer interval between dose 1 and 2 (≥31 days). After adjusting for dose 1 product and interval, the rate ratio for dose 2 mRNA-1273 compared to dose 2 BNT162b2 was 6.6 (95% CI 3.3-13.2) in males 18-24 who received their second dose on or after June 1, 2021.

**Table 3.** Reporting rate of myocarditis/pericarditis among males aged 18-24 years by vaccine products and inter-dose interval with dose 2 on or after June 1, 2021

| Vaccine schedule | Reports (N) | Doses administered (N) | Rate (95% CI) per million doses |
| --- | --- | --- | --- |
| People with two doses |  |  |  |
| BNT162b2-BNT162b2 | 11 | 235,819 | 46.6 (23.3 - 83.5) |
| Interval $\leq 30$ days | 2 | 21,160 | 94.5 (11.4 - 341.4) |
| Interval 31-55 days | 8 | 124,235 | 64.4 (27.8 - 126.9) |
| Interval $\geq 56$ days | 1 | 90,424 | 11.1 (0.3 - 61.6) |
| mRNA-1273-mRNA-1273 | 27 | 93,616 | 288.4 (190.1 - 419.6) |
| Interval $\leq 30$ days | 4 | 10,623 | 376.5 (102.6 - 964.1) |
| Interval 31-55 days | 20 | 60,352 | 331.4 (202.4 - 511.8) |
| Interval $\geq 56$ days | 3 | 22,641 | 132.5 (27.3 - 387.2) |
| mRNA-1273-BNT162b2 | 0 | 8,853 | 0 (0.0 - 416.7) |
| Interval $\leq 30$ days | 0 | 1,058 | 0.0 (0.0 - 3486.7) |
| Interval 31-55 days | 0 | 5,402 | 0.0 (0.0 - 682.9) |
| Interval $\geq 56$ days | 0 | 2,393 | 0.0 (0.0 - 1541.5) |
| BNT162b2-mRNA-1273 | 29 | 85,893 | 337.6 (226.1 - 484.9) |
| Interval $\leq 30$ days | 6 | 7,720 | 777.2 (285.2 - 1691.6) |
| Interval 31-55 days | 20 | 62,717 | 318.9 (194.8 - 492.5) |
| Interval $\geq 56$ days | 3 | 15,456 | 194.1 (40.0 - 567.2) |

**Figure 1.**
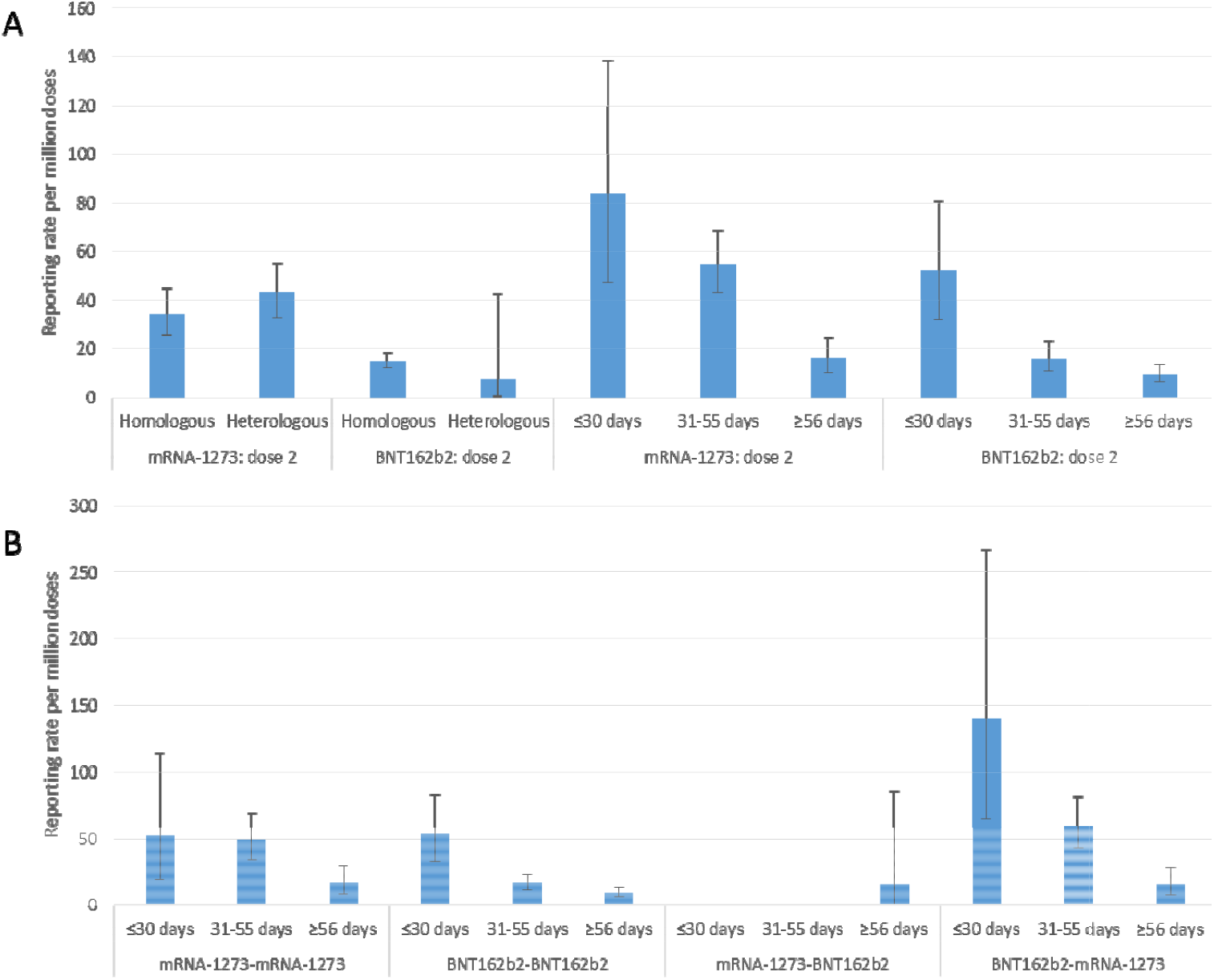
Overall reporting rate of myocarditis/pericarditis among people who have completed their two-dose series with dose 2 on or after June 1, 2021 by A) homologous/heterologous schedule and inter-dose interval and B) homologous/heterologous schedule by inter-dose interval

The number of observed events of myocarditis/pericarditis following vaccination exceeded that of the expected events based on a 7 day risk window for several age groups following dose 2, when restricted to dose 2 administration on/after June 1, 2021 (Table 4; analyses of the full period including dose 1 are provided in Supplementary Table 6). The ratio of observed to expected events was highest for males aged 18-24 following dose 2 of mRNA-1273 and for males aged 12-17 following dose 2 of BNT162b2.

**Table 4.** Observed vs. expected episodes of myocarditis/pericarditis using a 7-day risk window following dose 2 of COVID-19 mRNA vaccines among individuals receiving dose 2 on or after June 1, 2021, by age group, sex, and vaccine product

| Age group (years) | Females |  |  | Males |  |  |
| --- | --- | --- | --- | --- | --- | --- |
|  | Individuals with 2 doses | Expected* | Observed | Individuals with 2 doses | Expected* | Observed |
| **BNT162b2 – Dose 2** |  |  |  |  |  |  |
| 12-17 | 331,016 | 0.1-0.1 | **4** | 338,234 | 0.4-0.5 | **31** |
| 18-24 | 255,580 | 0.3-0.3 | **2** | 245,430 | 0.9-1.0 | **10** |
| 25-29 | 196,378 | 0.2-0.3 | **3** | 190,586 | 0.5-0.6 | **2** |
| 30-39 | 404,704 | 0.5-0.6 | **2** | 369,721 | 1.1-1.3 | **6** |
| 40-49 | 404,785 | 0.5-0.7 | 0 | 350,902 | 1.0-1.1 | **1** |
| 50-59 | 460,742 | 0.8-1.0 | 0 | 420,927 | 1.2-1.4 | **1** |
| 60-69 | 441,965 | 1.0-1.2 | 0 | 392,472 | 1.3-1.5 | **3** |
| 70-79 | 368,666 | 1.0-1.3 | 1 | 319,305 | 1.2-1.5 | **3** |
| ≥80 | 193,578 | 0.5-0.6 | 0 | 148,837 | 0.5-0.7 | 0 |
| **mRNA-1273 – Dose 2** |  |  |  |  |  |  |
| 12-17** | - | - | - | - | - | - |
| 18-24 | 170,317 | 0.2-0.2 | **7** | 179,866 | 0.6-0.7 | **55** |
| 25-29 | 133,420 | 0.1-0.2 | 0 | 151,079 | 0.4-0.5 | **12** |
| 30-39 | 266,347 | 0.3-0.4 | **5** | 292,548 | 0.9-1.0 | **15** |
| 40-49 | 261,699 | 0.4-0.4 | **2** | 274,340 | 0.8-0.9 | **5** |
| 50-59 | 292,890 | 0.5-0.6 | **1** | 311,910 | 0.9-1.0 | **2** |
| 60-69 | 247,723 | 0.6-0.7 | 0 | 249,489 | 0.8-0.9 | **2** |
| 70-79 | 139,124 | 0.4-0.5 | 0 | 128,971 | 0.5-0.6 | **1** |
| ≥80 | 66,729 | 0.2-0.2 | 0 | 47,684 | 0.2-0.2 | 0 |

\*The expected range is estimated from the confidence intervals around the mean background rate from 2015-2019.
\*\*Estimates were not provided for individuals aged 12-17 for mRNA-1273 because this product was not used for this age group in Ontario.
**Bold** results indicate where the observed number was greater than the upper confidence limit of the expected number.

## Discussion

Using passive vaccine safety surveillance data, we identified 297 reports of myocarditis/pericarditis following receipt of an mRNA vaccine that met the BC case definition since the start of the COVID-19 vaccine program in Ontario, Canada. Consistent with other surveillance systems and studies,^10,11^ we found that rates of myocarditis/pericarditis were highest among young males following dose 2, where they were tightly clustered within the first week after vaccination. Although rates were higher following a second dose of either mRNA vaccine as compared to a first dose, we observed a strong suggestion of a product-specific association; the rates following a second dose of mRNA-1273 were higher than those following a second dose of BNT162b2, in particular for young males. In addition to product specific insights for age/sex groups at highest risk, our analyses suggest that inter-dose interval and vaccine schedule combinations may also play a role in the risk of myocarditis/pericarditis. These observations suggest that there may be programmatic strategies relating to product, interval, and schedule that could play a role in reducing the risk of myocarditis/pericarditis following mRNA vaccines.

The crude reporting rates for myocarditis/pericarditis from Ontario are in line with estimates from other passive surveillance systems and other data sources,^2-5,12^ although there is variability in age-specific rates across systems and countries. In Israel, where only BNT162b2 vaccines were used following the product monograph with a 21 day inter-dose interval, the rate of myocarditis (using the BC definition) following dose two among males 16-19 was 150 per 1,000,000 between December 2020 and May 2021, although this time period encompassed both active and passive surveillance periods.^2^ The rate of myocarditis/pericarditis among males aged 12-17 who received two doses of BNT162b2 at an interval of 30 days or less in Ontario was similar at 159.7 per million doses. In the United Kingdom (UK), the reporting rate for myocarditis after both first and second doses across all ages was estimated at 10 per million doses of BNT162b2 and 36 per million doses of mRNA-1273 based on events submitted as of November 17, 2021.^4^ For those aged 19-29, rates of myocarditis following dose two were 22 and 69 per million for BNT162b2 and mRNA-1273, respectively. This trend of increased rate for mRNA-1273 is consistent with our findings, although the overall rate is lower. The UK employed an extended inter-dose interval and their overall results may be more comparable to our subgroup analyses reflecting the rates among individuals who had 8 or more weeks in between doses. Rates across data sources in the United States (US) vary. Using data from four FDA Biologics Effectiveness and SafeTy (BEST) administrative data claims databases, among males 18-25, the rate of myocarditis/pericarditis within 7 days following second dose mRNA-1273 ranged from 72.4 (95%CI 23.2-228.1) per million to 283.7 (95% CI 145.2-573.5) per million.^13^ In Ontario, we estimated a similar rate of myocarditis/pericarditis at 299.5 per million following a second dose of mRNA-1273 in males 18-24 years. Using data from the Vaccine Adverse Event Reporting System (VAERS), a passive reporting system, the reported rate of myocarditis per million doses in males within seven days of a second dose of mRNA-1273 was much lower than estimated in the BEST databases, with a rate of 38.5 per million.^14^ The rate per million following a second dose of BNT162b2 was 36.8 in males aged 18-24 years, 69.1 in those aged 16-17 years and 39.9 in those aged 12-15 years.^14^ Data from the US also include those from the active surveillance system, Vaccine Safety Datalink (VSD), with reporting rates higher than in VAERS.^15^ In a head-to-head analysis of BNT162b2 and mRNA-1273 among those aged 18-39 years, the VSD reported the adjusted rate of myocarditis/pericarditis within 7 days of dose two was 2.72 times greater (95% CI 1.25-6.05) for those who received mRNA-1273 as compared to BNT162b2, with an excess of 13.3 cases per million second doses of mRNA-1273 vs. BNT162b2.^16^ There are several possible explanations for differences in reporting rates across systems, including outcomes studied (i.e., myocarditis only versus myocarditis/pericarditis), different case definitions used to classify outcomes, completeness in reporting, and health system context (i.e., access to publicly-funded health services). Finally, our analyses suggest that country-specific differences in the inter-dose interval and heterologous schedules may be an additional influence on variability in reporting rates across jurisdictions.

Following extensive review and discussion of the product-specific differences identified from passive vaccine safety surveillance, Ontario modified its COVID-19 vaccine program on September 29, 2021 to preferentially offer the BNT162b2 vaccine to individuals 12-24 years of age.^7^ Although authorized by Health Canada for adolescents 12-17 years of age in late August 2021, the mRNA-1273 vaccine has yet to be incorporated into Ontario’s adolescent vaccine program. As of mid-November 2021, several countries, including Norway, Sweden, Finland, France, and Germany, have issued similar guidance limiting the use of mRNA-1273 in those adolescents and young adults.^17-20^

Although data on the possible relative risks between products for myocarditis/pericarditis are emerging, these findings need to be considered within the context of absolute risk, as myocarditis/pericarditis is still a rare or very rare event, based on standard pharmacovigilance definitions.^21^ Importantly, the risk of myocarditis/pericarditis following mRNA vaccines also needs to be considered in relation to risks of myocarditis following SARS-CoV-2 infection (i.e., higher rates of myocarditis following infection than vaccination)^22-24^ and the high effectiveness of mRNA vaccine products, including some suggestion of a more durable response following mRNA-1273 vaccine.^25^

These analyses include data on all AEFI entered into a single passive vaccine safety surveillance system in a large jurisdiction with high vaccine coverage (77.6% two-dose coverage among the vaccine eligible population [i.e., ≥12 years of age] as of September 4, 2021). All AEFI reports were individually reviewed by a team of specialized nurses and physicians to limit analyses to those events meeting BC case definitions for myocarditis or pericarditis (levels 1-3). We utilized data on the entire vaccine program through the provincial COVID-19 vaccine registry, which allowed us to examine reporting rates in the context of detailed denominator data relating to various product schedules and intervals. Lastly, we used historical data from the same population giving rise to these outcomes, in a jurisdiction with universal access to publicly-funded health services, which allowed for the comparison of observed versus expected events in the context of the vaccination program. Despite these strengths, there are several limitations in this analysis worth noting, including those inherent to passive vaccine safety surveillance systems such as stimulated reporting during the period of enhanced reporting. However, these limitations were minimized by a restriction of events only to those meeting BC levels 1-3, and thorough sensitivity analyses; when we analyzed our rates in different time periods, as well as restricted our analysis to myocarditis only (BC levels 1-2), our conclusions were unchanged. Lastly, several of our reporting rates for product and schedule combinations were based on small numbers, leading to very wide confidence intervals; as such, rates for individual strata should be interpreted with caution.

## Conclusions

Although myocarditis/pericarditis following mRNA vaccines is rare, our analyses suggest that modifications to mRNA COVID-19 vaccine programs relating to age-based product considerations and the use of longer inter-dose intervals may reduce the risk of these events. Confirmation of these findings, and further exploration of the influence of heterologous mRNA vaccine schedules on the risk of myocarditis/pericarditis, are needed.

## Data Availability

Public Health Ontario (PHO) cannot disclose the underlying data. Doing so would compromise individual privacy contrary to PHOs ethical and legal obligations. Restricted access to the data may be available under conditions prescribed by the Ontario Personal Health Information Protection Act, 2004, the Ontario Freedom of Information and Protection of Privacy Act, the Tri-Council Policy Statement: Ethical Conduct for Research Involving Humans (TCPS 2 (2018)), and PHO privacy and ethics policies. Data are available for researchers who meet PHOs criteria for access to confidential data.  Information about PHOs data access request process is available on-line at https://www.publichealthontario.ca/en/data-and-analysis/using-data/data-requests.

## Acknowledgments

The authors thank Lennon Li, Lauren Paul, and Leigh Hobbs for their contributions to the data analysis.

## Funding

This work was supported by Public Health Ontario. This work was also supported by the Canadian Immunization Research Network (CIRN) through a grant from the Public Health Agency of Canada and the Canadian Institutes of Health Research (CNF 151944). Additionally, this work was supported by ICES, which is funded by an annual grant from the Ontario Ministry of Health (MOH). JCK is supported by a Clinician-Scientist Award from the University of Toronto Department of Family and Community Medicine. The analyses, conclusions, opinions and statements expressed herein are solely those of the authors and do not reflect those of the funding or data sources; no endorsement is intended or should be inferred.

## Conflict of interest

The authors declare no conflicts of interest.

## Ethics approval

The Public Health Ontario Ethics Review Board has determined that this project did not require research ethics committee approval as the activities described in this manuscript were conducted in fulfillment of Public Health Ontario’s legislated mandate “to provide scientific and technical advice and support to the health care system and the Government of Ontario in order to protect and promote the health of Ontarians” (Ontario Agency for Health Protection and Promotion Act, SO 2007, c 10) and are therefore considered public health practice, not research.

## Notes

### Competing Interest Statement

The authors have declared no competing interest.

### Author Declarations

The Public Health Ontario Ethics Review Board has determined that this project did not require research ethics committee approval as the activities described in this manuscript were conducted in fulfillment of Public Health Ontarios legislated mandate to provide scientific and technical advice and support to the health care system and the Government of Ontario in order to protect and promote the health of Ontarians (Ontario Agency for Health Protection and Promotion Act, SO 2007, c 10) and are therefore considered public health practice, not research.

