## Supplementary materials for "Epidemiology of myocarditis and pericarditis following mRNA vaccines in Ontario, Canada: by vaccine product, schedule and interval"

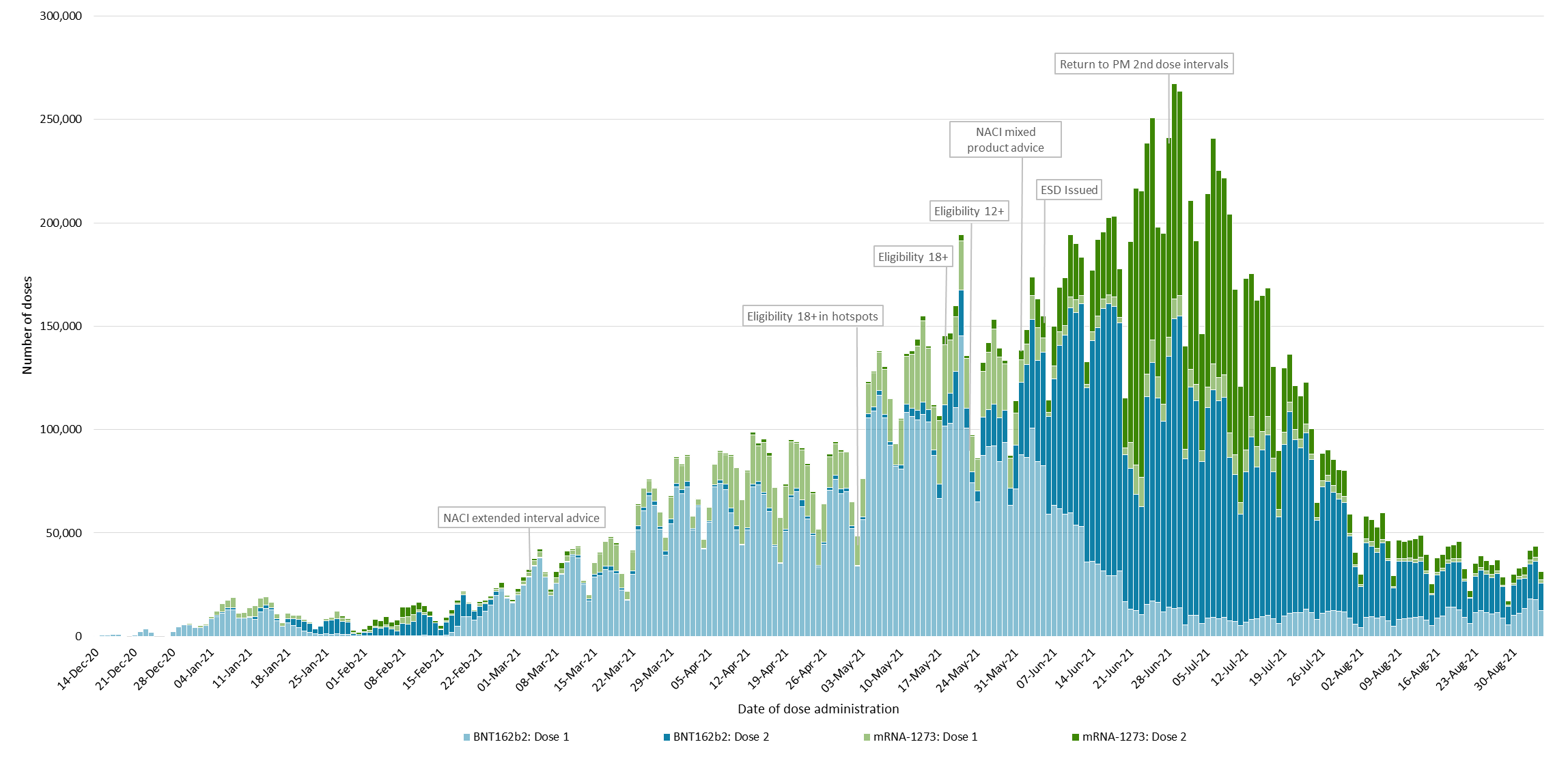
**Figure 1**. Number of COVID-19 mRNA vaccine doses administered in Ontario by dose number and vaccine product and overview of vaccine program changes

NACI: National Advisory Committee on Immunization; ESD: Enhanced Surveillance Directive; PM: Product Monograph

**Figure 2.** Myocarditis/pericarditis reports following COVID-19 mRNA vaccines by dose number and time to symptom onset*

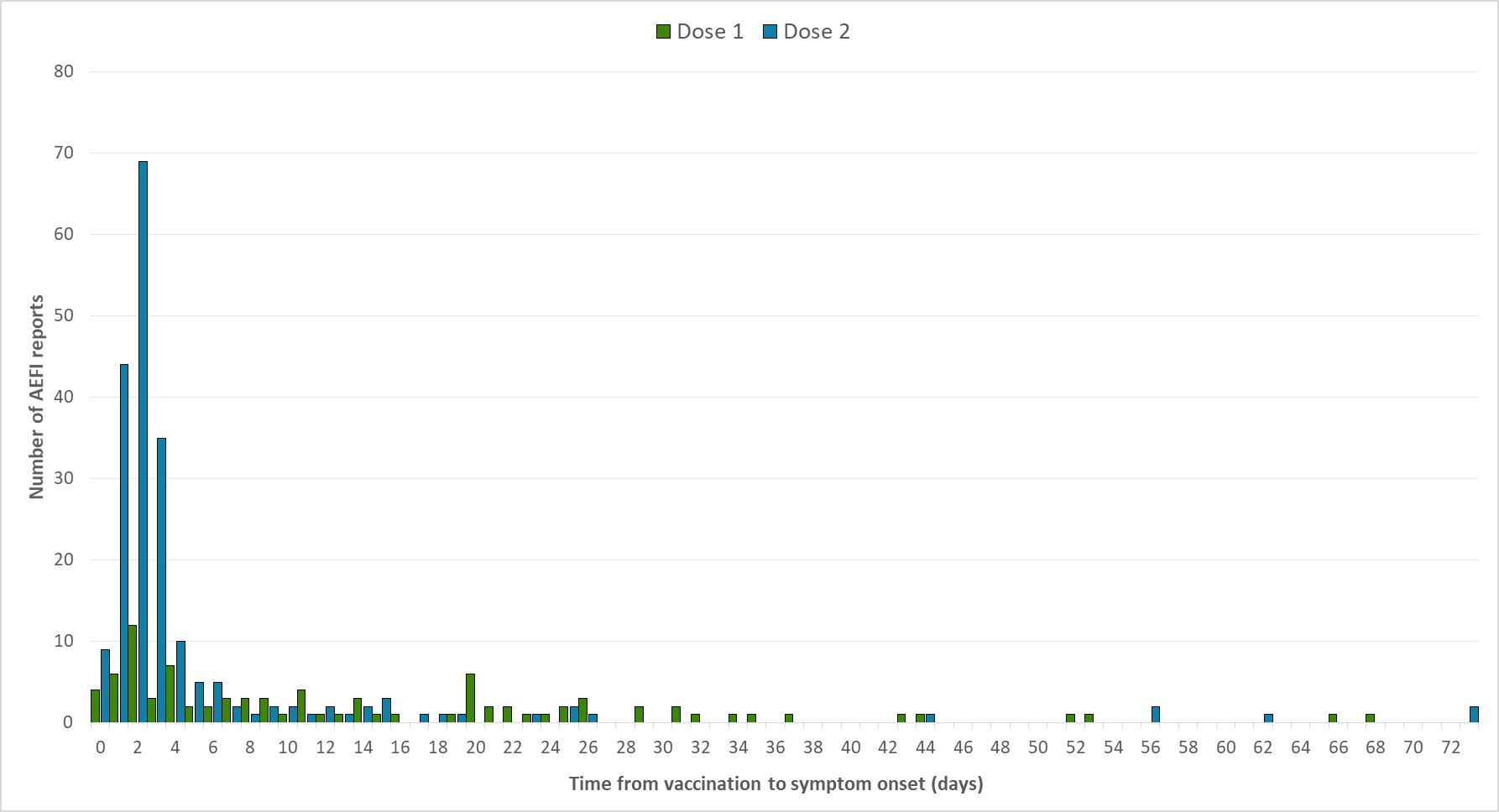

*2 reports with unknown time to onset were excluded from this figure

**Table 1**. Definition for identifying myocarditis/pericarditis using administrative data to estimate background rates^1^

| **Outcome** | **Definition** |
| --- | --- |
| **Myocarditis/pericarditis** | Acute pericarditis, pericarditis in disease classified elsewhere, acute myocarditis, myocarditis in diseases classified elsewhere, myocarditis, unspecified:  ICD-10-CA codes: I30.x, I32.x, I40.x, I41.x, I51.4 |

**Table 2.** Clinical diagnosis and severity of myocarditis/pericarditis

|  | Myocarditis (n=105) | Pericarditis (n=85) | Myopericarditis* (n=107) | Total (N=297) |
| --- | --- | --- | --- | --- |
| Age group (years) |  |  |  |  |
| 12-17 | 28 (26.7%) | 8 (9.4%) | 19 (17.8%) | 55 (18.5%) |
| 18-24 | 30 (28.6%) | 16 (18.8%) | 50 (46.7%) | 96 (32.3%) |
| 25-39 | 30 (28.6%) | 12 (14.1%) | 30 (28.0%) | 72 (24.2%) |
| ≥40 | 17 (16.2%) | 49 (57.6%) | 8 (7.5%) | 74 (24.9%) |
| Healthcare utilization/outcome |  |  |  |  |
| Emergency department | 101 (96.2%) | 83 (97.6%) | 106 (99.1%) | 290 (97.6%) |
| In-patient hospitalization | 87 (82.9%) | 33 (38.8%) | 90 (84.1%) | 210 (70.7%) |
| Intensive care unit admission | 5 (4.8%) | 5 (5.9%) | 4 (3.7%) | 14 (4.7%) |
| Death | 0 | 0 | 0 | 0 |
| BC level |  |  |  |  |
| 1 | 34 (32.4%) | 5 (5.9%) | 28 (26.2%) | 67 (22.6%) |
| 2 | 68 (64.8%) | 73 (85.9%) | 79 (73.8%) | 220 (74.1%) |
| 3 | 3 (2.9%) | 7 (8.2%) | 0 (0.0%) | 10 (3.4%) |

*Myopericarditis reports were assessed against Brighton Collaboration (BC) case definitions for myocarditis and pericarditis. The highest level of certainty was assigned to the report (e.g., if a report met level 1 for myocarditis and level 3 for pericarditis, it was assigned as level 1).

**Table 3.** Crude reporting rate of myocarditis meeting BC level 1 and 2 only per million doses administered by vaccine product, dose number, age, and sex: series initiated on or after June 1, 2021

| BNT162b2 | | | | | | |
| --- | --- | --- | --- | --- | --- | --- |
|  | All | | Female | | Male | |
| Age group (years) | Dose 1 | Dose 2 | Dose 1 | Dose 2 | Dose 1 | Dose 2 |
| 12-17 | 21.5 (10.7 - 38.4) | 49.7 (30.8 – 76.0) | 8.1 (1.0 - 29.1) | 9.7 (1.2 - 35.1) | 34.2 (15.6 - 64.9) | 88.1 (53.0 - 137.5) |
| 18-24 | 10.7 (2.2 - 31.3) | 19.0 (3.9 - 55.5) | 7.9 (0.2 - 44.1) | 0.0 (0.0 - 50.5) | 13.1 (1.6 - 47.3) | 35.5 (7.3 - 103.7) |
| 25-39 | 9.3 (3.0 - 21.7) | 12.8 (3.5 - 32.9) | 0.0 (0.0 - 14.3) | 13.1 (1.6 - 47.5) | 17.9 (5.8 - 41.8) | 12.6 (1.5 - 45.4) |
| ≥40 | 0.0 (0.0 - 7.3) | 0.0 (0.0 - 11.7) | 0.0 (0.0 - 14.8) | 0.0 (0.0 - 23.5) | 0.0 (0.0 - 14.4) | 0.0 (0.0 - 23.3) |
| Total | 10.2 (6.1 - 15.9) | 23.2 (15.4 - 33.5) | 3.4 (0.7 - 9.8) | 6.8 (1.9 - 17.4) | 16.6 (9.5 – 27.0) | 38.8 (24.9 - 57.8) |
| mRNA-1273 | | | | | | |
|  | All | | Female | | Male | |
| Age group (years) | Dose 1 | Dose 2 | Dose 1 | Dose 2 | Dose 1 | Dose 2 |
| 12-17* | - | - | - | - | - | - |
| 18-24 | 0.0 (0.0 - 39.8) | 195.5 (117.7 - 305.3) | 0.0 (0.0 - 95.1) | 69.1 (14.2 - 201.9) | 0.0 (0.0 - 68.7) | 299.5 (171.2 - 486.4) |
| 25-39 | 16.2 (3.3 - 47.3) | 48.9 (23.5 – 90.0) | 0.0 (0.0 - 45.4) | 21.5 (2.6 - 77.7) | 28.8 (5.9 - 84.3) | 72.1 (31.1 – 142.0) |
| ≥40 | 10.0 (1.2 - 36.1) | 0.0 (0.0 – 19.0) | 0.0 (0.0 - 40.5) | 0.0 (0.0 - 40.9) | 18.3 (2.2 - 66.2) | 0.0 (0.0 - 35.6) |
| Total | 10.4 (3.4 - 24.4) | 58.4 (39.1 - 83.9) | 0.0 (0.0 - 17.5) | 22.0 (7.1 - 51.4) | 18.7 (6.1 - 43.7) | 89.4 (57.3 – 133.0) |

*Estimates were not provided for individuals aged 12-17 for mRNA-1273 because this product was not used for this age group in Ontario.

**Table 4.** Crude reporting rate of myocarditis/pericarditis per million doses administered by vaccine product, dose number, age, and sex: series initiated on or after December 14, 2020

| BNT162b2 | | | | | | |
| --- | --- | --- | --- | --- | --- | --- |
|  | All | | Female | | Male | |
| Age group (years) | Dose 1 | Dose 2 | Dose 1 | Dose 2 | Dose 1 | Dose 2 |
| 12-17 | 24.7 (14.9 - 38.6) | 53.6 (37.5 - 74.2) | 15.9 (5.8 - 34.6) | 12.0 (3.3 - 30.8) | 33.4 (17.8 – 57.0) | 94.5 (64.6 - 133.4) |
| 18-24 | 18.8 (10.5 - 31.1) | 24.3 (13.0 - 41.6) | 7.5 (1.5 - 21.8) | 7.2 (0.9 - 25.9) | 30.6 (15.8 - 53.5) | 43.4 (21.7 - 77.6) |
| 25-39 | 8.2 (4.6 - 13.4) | 13.2 (7.7 - 21.2) | 5.3 (1.7 - 12.3) | 11.7 (5.0 – 23.0) | 11.3 (5.4 - 20.7) | 15.0 (6.9 - 28.5) |
| ≥40 | 4.2 (2.5 - 6.5) | 6.4 (4.2 - 9.5) | 2.0 (0.7 - 4.7) | 4.7 (2.3 - 8.7) | 6.8 (3.7 - 11.3) | 8.5 (4.8 - 14.1) |
| Total | 8.5 (6.6 - 10.8) | 14.3 (11.5 - 17.5) | 4.5 (2.7 - 7.1) | 7.0 (4.5 - 10.5) | 13.1 (9.7 - 17.3) | 22.7 (17.6 - 28.8) |
| mRNA-1273 | | | | | | |
|  | All | | Female | | Male | |
| Age group (years) | Dose 1 | Dose 2 | Dose 1 | Dose 2 | Dose 1 | Dose 2 |
| 12-17 | - | - | - | - | - | - |
| 18-24 | 16.2 (4.4 - 41.4) | 177.8 (136.9 - 227.1) | 17.4 (2.1 – 63.0) | 45.7 (19.7 – 90.0) | 15.2 (1.8 - 54.8) | 305.2 (230.6 - 396.4) |
| 25-39 | 10.5 (3.9 - 22.9) | 39.1 (27.1 - 54.6) | 7.5 (0.9 - 27.1) | 12.1 (3.9 - 28.2) | 13.3 (3.6 - 33.9) | 63.9 (42.8 - 91.8) |
| ≥40 | 10.1 (5.2 - 17.7) | 8.3 (4.9 - 13.2) | 8.2 (2.7 - 19.1) | 3.7 (1.0 - 9.4) | 12.3 (4.9 - 25.3) | 13.2 (7.2 - 22.1) |
| Total | 11.0 (6.9 - 16.6) | 34.2 (28.3 - 41) | 9.1 (4.1 - 17.2) | 10.1 (5.9 - 16.2) | 12.9 (6.9 - 22.1) | 58.2 (47.3 - 70.9) |

*Estimates were not provided for individuals aged 12-17 for mRNA-1273 because this product was not used for this age group in Ontario.

**Figure 3.** Reporting rate following dose 2 of mRNA vaccine for all sexes in Ontario as of September 4, 2021

Reporting rate per million doses administered

| Age (years) | BNT162b2 | mRNA-1273 |
| --- | --- | --- |
| 12 | 26.6 | 0 |
| 13 | 73.1 | 0 |
| 14 | 54 | 0 |
| 15 | 35.8 | 0 |
| 16 | 70.1 | 0 |
| 17 | 61.8 | 0 |
| 18 | 70.5 | 262.8 |
| 19 | 28.1 | 230.5 |
| 20 | 13.6 | 197.6 |
| 21 | 26.3 | 228.1 |
| 22 | 13 | 152.7 |
| 23 | 12.2 | 110 |
| 24 | 11.9 | 88.7 |
| 25 | 11.6 | 34.5 |
| 26 | 23 | 50.6 |
| 27 | 0 | 51.5 |
| 28 | 11.7 | 51.2 |
| 29 | 23.6 | 33.9 |
| 30 | 34.8 | 67.3 |
| 31 | 0 | 16.7 |
| 32 | 23.3 | 0 |
| 33 | 11.8 | 17.7 |
| 34 | 35.4 | 70 |
| 35 | 0 | 34.7 |
| 36 | 0 | 52 |
| 37 | 0 | 52.9 |
| 38 | 11.6 | 52.4 |
| 39 | 11.8 | 0 |
| 40 | 0 | 35.3 |
| 41 | 0 | 36.4 |
| 42 | 12.2 | 18.4 |
| 43 | 0 | 0 |
| 44 | 0 | 0 |
| 45 | 11.9 | 0 |
| 46 | 0 | 36.2 |
| 47 | 0 | 0 |
| 48 | 0 | 0 |
| 49 | 0 | 0 |
| 50 | 0 | 16.5 |
| 51 | 0 | 0 |
| 52 | 21.5 | 0 |
| 53 | 0 | 33.5 |
| 54 | 0 | 16.6 |
| 55 | 0 | 15.9 |
| 56 | 9.6 | 0 |
| 57 | 9.5 | 14.7 |
| 58 | 9.5 | 0 |
| 59 | 9.9 | 15.9 |
| 60 | 0 | 0 |
| 61 | 21.5 | 0 |
| 62 | 0 | 0 |
| 63 | 11.3 | 18.5 |
| 64 | 0 | 0 |
| 65 | 32.2 | 38.4 |
| 66 | 10.8 | 0 |
| 67 | 0 | 0 |
| 68 | 11.6 | 0 |
| 69 | 11.4 | 0 |
| 70 | 11.5 | 0 |
| 71 | 0 | 0 |
| 72 | 11.9 | 0 |
| 73 | 0 | 0 |
| 74 | 11.7 | 30.3 |
| 75 | 0 | 0 |
| 76 | 0 | 0 |
| 77 | 16.1 | 0 |
| 78 | 0 | 0 |
| 79 | 37.3 | 0 |
| 80 | 0 | 0 |
| 81 | 43 | 0 |
| 82 | 0 | 0 |
| 83 | 0 | 0 |
| 84 | 0 | 0 |
| 85 | 0 | 0 |
| 86 | 0 | 0 |
| 87 | 0 | 0 |
| 88 | 0 | 0 |
| 89 | 0 | 0 |
| 90 | 0 | 0 |

**Table 5.** Rate or myocarditis/pericarditis per million by dose 2 product and interval, for individuals receiving dose 2 on or after June 1, 2021

| **BNT162b2 – Dose 2** | | | | | | | |
| --- | --- | --- | --- | --- | --- | --- | --- |
|  | Females | Males | 12-17 years** | 18-24 years | 25-39 years | ≥40 years | OVERALL |
| **Product schedule*** |  |  |  |  |  |  |  |
| BNT162b2- BNT162b2 | 5.9 (3.5 – 9.5) | 25.0 (19.3 – 32.0) | 53.8 (37.7 - 74.5) | 26.9 (14.3 - 45.9) | 13.4 (7.5 - 22.1) | 5.4 (3.1 - 8.6) | 14.9 (11.9 - 18.6) |
| mRNA-1273 –BNT162b2 | 0.0 (0.0 - 58.5) | 14.6 (0.4 - 81.2) | - | 0.0 (0.0 - 218.8) | 0.0 (0.0– 107.0) | 12.5 (0.3 - 69.7) | 7.6 (0.2 - 42.3) |
| **Interval (days)** |  |  |  |  |  |  |  |
| ≤30 | 25.8 (8.4 - 60.3) | 79.3 (44.4 - 130.8) | 101.9 (55.7 - 170.9) | 45.3 (5.5 - 163.7) | 42.5 (11.6 - 108.7) | 0.0 (0.0 – 34.4) | 52.1 (31.8 - 80.5) |
| 31-55 | 4.1 (1.1 - 10.6) | 28.2 (18.6 – 41.0) | 37.7 (21.6 - 61.3) | 34.7 (15.9 - 66) | 8.7 (2.8 - 20.3) | 1.5 (0.0 - 8.3) | 16.1 (10.9 - 22.8) |
| ≥56 | 4.5 (2.0 – 9.0) | 15.6 (9.9 - 23.4) | 55.7 (20.4 - 121.2) | 10.1 (1.2 - 36.5) | 12.3 (4.5 - 26.7) | 6.9 (4.0 - 11.1) | 9.6 (6.5 - 13.6) |
| **mRNA-1273 – Dose 2** | | | | | | | |
|  | Females | Males | 12-17 years** | 18-24 years | 25-39 years | ≥40 years | OVERALL |
| **Product schedule** |  |  |  |  |  |  |  |
| mRNA-1273 -mRNA-1273 | 9.5 (3.8 - 19.6) | 58.3 (42.4 - 78.3) | - | 162.0 (108.5 - 232.6) | 30.1 (16.0 - 51.4) | 10.2 (4.7 - 19.4) | 34.2 (25.4 - 44.9) |
| BNT162b2 -mRNA-1273 | 13.1 (6.0 – 25.0) | 72.5 (53.8 - 95.6) | - | 203.9 (142.0 - 283.6) | 52.0 (32.2 - 79.5) | 3.8 (0.8 – 11.0) | 42.9 (32.6 - 55.3) |
| **Interval (days)** |  |  |  |  |  |  |  |
| ≤30 | 36.2 (7.5 - 105.8) | 125.7 (64.9 - 219.5) | - | 353.1 (182.4 - 616.8) | 39.5 (8.1 - 115.4) | 0.0 (0.0 – 53.9) | 83.9 (47.0 - 138.4) |
| 31-55 | 9.4 (3.5 - 20.5) | 96.0 (74.4 - 121.9) | - | 184.0 (133.7 – 247.0) | 45.0 (29.1 - 66.4) | 7.4 (2.0 – 19.0) | 54.6 (42.8 - 68.7) |
| ≥56 | 10.0 (4.0 - 20.6) | 23.1 (12.9 – 38.0) | - | 103.2 (44.5 - 203.3) | 29.4 (10.8 - 64) | 7.5 (3.2 - 14.7) | 16.2 (10.2 - 24.6) |

*There were 5 reports following ChAdOx1-mRNA-1273 among 350,628 doses administered and 5 reports following ChAdOx1-BNT162b2 among 279,805 doses administered. These are excluded from Table 5.

**Estimates were not provided for individuals aged 12-17 for mRNA-1273 because this product was not used for this age group in Ontario.

**Table 6.** Observed vs. expected episodes of myocarditis/pericarditis using a 7 day risk window following COVID-19 mRNA vaccines, by dose number, age group, sex, and vaccine product: series initiated on or after December 14, 2020

| Age group (years) | Females | | | Males | | |
| --- | --- | --- | --- | --- | --- | --- |
|  | Individuals with 1 dose | Expected* | Observed | Individuals with 1 dose | Expected* | Observed |
| **BNT162b2 – Dose 1** | | | | | | |
| 12-17 | 379,140 | 0.1-0.2 | **2** | 391,153 | 0.5-0.6 | **9** |
| 18-24 | 404,251 | 0.4-0.5 | **2** | 394,006 | 1.4-1.6 | **8** |
| 25-29 | 320,290 | 0.3-0.4 | 0 | 311,179 | 0.9-1.1 | 1 |
| 30-39 | 630,243 | 0.7-0.9 | **2** | 581,517 | 1.7-2.0 | **3** |
| 40-49 | 556,678 | 0.8-0.9 | 0 | 461,753 | 1.3-1.5 | 1 |
| 50-59 | 610,789 | 1.1-1.3 | 1 | 529,436 | 1.5-1.7 | 0 |
| 60-69 | 562,757 | 1.3-1.5 | 0 | 488,803 | 1.6-1.8 | 0 |
| 70-79 | 459,321 | 1.3-1.6 | 0 | 398,875 | 1.6-1.9 | 0 |
| ≥80 | 274,282 | 0.7-0.9 | 0 | 198,072 | 0.7-0.9 | 0 |
| **mRNA-1273 – Dose 1** | | | | | | |
| 12-17** | - | - | - | - | - | - |
| 18-24 | 115,633 | 0.1-0.2 | 0 | 132,730 | 0.5-0.5 | **1** |
| 25-29 | 90,224 | 0.1-0.1 | 0 | 107,662 | 0.3-0.4 | **3** |
| 30-39 | 177,843 | 0.2-0.3 | **1** | 196,422 | 0.6-0.7 | 0 |
| 40-49 | 145,018 | 0.2-0.2 | **1** | 148,715 | 0.4-0.5 | **2** |
| 50-59 | 153,080 | 0.3-0.3 | 0 | 158,275 | 0.4-0.5 | **1** |
| 60-69 | 143,787 | 0.3-0.4 | 0 | 141,062 | 0.5-0.5 | 0 |
| 70-79 | 88,857 | 0.3-0.3 | **1** | 80,904 | 0.3-0.4 | 0 |
| ≥80 | 82,842 | 0.2-0.3 | 0 | 45,978 | 0.2-0.2 | 0 |
| **BNT162b2 – Dose 2** | | | | | | |
| 12-17 | 332,911 | 0.1-0.1 | **4** | 339,559 | 0.4-0.6 | **31** |
| 18-24 | 280,256 | 0.3-0.4 | **2** | 254,608 | 0.9-1.0 | **10** |
| 25-29 | 225,758 | 0.2-0.3 | **3** | 202,924 | 0.6-0.7 | **2** |
| 30-39 | 461,226 | 0.5-0.7 | **4** | 398,150 | 1.2-1.3 | **6** |
| 40-49 | 465,951 | 0.6-0.8 | 0 | 378,631 | 1.0-1.2 | 1 |
| 50-59 | 530,800 | 0.9-1.1 | 0 | 453,440 | 1.3-1.5 | 1 |
| 60-69 | 488,865 | 1.1-1.3 | 0 | 420,538 | 1.3-1.6 | **3** |
| 70-79 | 393,725 | 1.1-1.3 | 1 | 340,235 | 1.3-1.6 | **3** |
| ≥80 | 242,747 | 0.6-0.8 | 0 | 174,289 | 0.6-0.8 | 0 |
| **mRNA-1273 – Dose 2** | | | | | | |
| 12-17** | - | - | - | - | - | - |
| 18-24 | 175,568 | 0.2-0.2 | **7** | 183,837 | 0.7-0.8 | **55** |
| 25-29 | 138,273 | 0.1-0.2 | 0 | 154,706 | 0.4-0.5 | **12** |
| 30-39 | 276,662 | 0.3-0.4 | **5** | 299,961 | 0.9-1.0 | **15** |
| 40-49 | 273,693 | 0.4-0.5 | **2** | 282,300 | 0.8-0.9 | **5** |
| 50-59 | 307,704 | 0.5-0.6 | **1** | 321,942 | 0.9-1.0 | **3** |
| 60-69 | 261,343 | 0.6-0.7 | 0 | 260,313 | 0.8-1.0 | **2** |
| 70-79 | 150,699 | 0.4-0.5 | 0 | 138,649 | 0.5-0.6 | **1** |
| ≥80 | 104,124 | 0.3-0.3 | 0 | 62,645 | 0.2-0.3 | 0 |

*The expected range is estimated from the confidence intervals around the mean background rate from 2015-2019.

**Estimates were not provided for individuals aged 12-17 for mRNA-1273 because this product was not used for this age group in Ontario.

**Bold** results indicate where the observed number was greater than the upper confidence limit of the expected number.
